# Molecular point-of-care testing for SARS-CoV-2 using the ID NOW™ System in Emergency Department: Prospective Evaluation and Implementation in the Care Process

**DOI:** 10.1101/2021.09.09.21263266

**Authors:** S. Kortüm, M. Krause, H.-J. Ott, L. Kortüm, H-P. Schlaudt

## Abstract

**Background:** The increasing number of cases and hospital admissions due to COVID-19 created an urgent need for rapid, reliable testing procedures for SARS-CoV-2 in Emergency Departments (ED) in order to effectively manage hospital resources, allocate beds and prevent nosocomial spread of infection. The ID NOW™ COVID-19 assay is a simple, user-friendly, rapid molecular test run on an instrument with a small footprint enabling point-of-care diagnostics.

**Methods:** In the first wave, outsourced RT-PCR testing regularly required 36-48 hours before results were available. This prospective study was conducted in the second wave (October 2020-April 2021) and evaluated the impact the implementation of the ID NOW™ COVID-19 test in the ED had on clinical care processes and patient pathways. 710 patients were recruited upon arrival at the ED which included those presenting clinical symptoms, asymptomatic individuals or persons fulfilling epidemiological criteria. The first anterior nasal swab was taken by trained nurses in the ambulance or a separate consultation room. The ID NOW™ COVID-19 test was performed in the ED in strict compliance with the manufacturer’s instructions and positive or suspected cases were additionally tested with RT_PCR (cobas SARS-COV-2 RT-PCR, Roche) following collection of a second nasopharyngeal NP specimen.

**Results:** Swabs directly tested with the ID NOW™ COVID-19 test showed a diagnostic concordance of 98 % (sensitivity 99.59 %, specificity 94.55 %, PPV 97.6 %, NPV 99.05 %) compared to RT-PCR as reference. The 488 patients that tested positive with the ID NOW™ COVID-19 had a Ct range in RT-PCR results between 7.94 to 37.42 (in 23.2 % > 30). Two false negative results (0.28%) were recorded from patients with Ct values > 30. 14 (1.69%) discordant results were reviewed case-by-case and usually associated with either very early or very advanced stages of infection. Furthermore, patients initially negative with the ID NOW™ COVID-19 test and admitted to the hospital were tested again on days 5 and 12: no patient became positive.

**Discussion:** The ID NOW™ COVID-19 test for detection of SARS-CoV-2 demonstrated excellent diagnostic agreement with RT-PCR under the above-mentioned patients pathways implemented during the second wave. The main advantage of the system was the provision of reliable results within a few minutes. This not only allowed immediate initiative of appropriate therapy and care for COVID-19 (patient benefit) but provided essential information on isolation and thus available beds. This drastically helped the overall finances of the department and additionally allowed more patients to be admitted including those requiring immediate attention; this was not possible during the first wave since beds were blocked waiting for diagnostic confirmation. Our findings also show that when interpreting the results, the clinical condition and epidemiological history of the patient must be taken into account, as with any test procedure. Overall, the ID NOW™ COVID-19 test for SARS-CoV-2 provided a rapid and reliable alternative to laboratory-based RT-PCR in the real clinical setting which became an acceptable part of the daily routine within the ED and demonstrated that early patient management can mitigate the impact of the pandemic on the hospital.

## Introduction

The SARS-CoV-2 pandemic has been ongoing since January 2020 and identifying patients with COVID-19 as quickly as possible has been essential for maintaining and optimising care processes in emergency departments. A timely accurate classification of patients into infectious vs. non-infectious is essential to avoid nosocomial infections with SARS-CoV-2. Moreover, it enables acceleration of patient admission as well as a rapid and targeted allocation of patients to appropriate treatment areas [1, 2].

The Klinikum Hochrhein, has an isolated inpatient area with an intake capacity of up to 46 patients and is located in part of the building with its own access from the outside and thus, can be easily segregated from the main building. This allows patient pathways (defined as overall care) to be easily designated upon arrival of the individuals. This separation was highly advantageous during the pandemic since it provided an immediate solution for isolation. If demands exceeded the existing isolation capacities, inpatient areas in the main building would have been reallocated but then would have no longer been available for regular care.

Experiences from the so-called “first wave” of the pandemic in the area surrounding the clinic (February to May 2020), highlighted that a correct classification of patients is not possible when solely based on clinical and epidemiological criteria [3]. Real-time PCR has been considered the gold standard in the diagnosis of SARS-CoV-2 infections but results take at least 24 hours (on average 36-48 hours) at the Klinikum Hochrhein since this testing has to be outsourced to an external laboratory. This is actually common practice since most hospitals do not have their own in-house facilities. During the initial stage of the pandemic, the application of the clinical-epidemiological algorithms, developed by the Klinikum Hochrhein, led to the isolation of a large number of patients over at least 24 hours as a precaution even though only 20% were actually confirmed as a “positive case” [3]. Against a rising tide of incidences and hospital admissions from Autumn 2020 onwards, there was an urgent need for rapid, reliable testing procedures for detecting cases of SARS-CoV-2 in our Emergency Department (emergency care level 2). This was essential since there was an urgent need to allocate beds in a targeted manner, to pool infectious patients as early as possible and ultimately enable inpatient capacities to be maintained for regular care outside the pandemic. Thus, we employed the isothermal nucleic acid amplification-based ID NOW™ COVID-19 assay from Abbotts which is a simple, user-friendly rapid molecular NAAT (Nucleic Acid Amplification Technology) test that does not require specific microbiological equipment or laboratory space. This point-of-care diagnostic device is a molecular isothermal test based on NEAR (nicking endonuclease amplification reaction) technology, and detects the RdRP gene of SARS-CoV-2 with a manufacturer’s specified limit of detection (LOD) of 125 genome equivalents/ml [4]. The test results are available after approx. 13 minutes.

Results reported in the literature on the reliability of the system have been variable [5-9]. However, a recent study and review from Farfour et al., showed that when data were assessed on the basis of CT-values (≤35) sensitivity reflected the outcome via PCR [10]. There is a correlation between how the samples are collected or manipulated and compliance with the manufacturer’s instructions for performing the test [2, 10, 11]. In particular, dissolution of the sample in a transport medium (VTM) and delayed processing of the sample material (> 2 hour) significantly impairs the sensitivity of the test [1, 2, 12]. This was recently demonstrated in an ER setting by Nguyen Van and colleagues using nasopharyngeal (NP) swabs since sensitivity dropped 98.0% to 62.5% when samples were placed in VTM solution and measured 3 hours after collection [2].

The purpose of this work was to evaluate the ID NOW™ COVID-19 testing platform in comparison to RT-PCR under real clinical conditions after implementation of the device into the daily workflow and processes in an emergency department with the aim of reliably identifying incoming patients with early stages of infection so that informed decisions regarding isolation or infectious patients could be swiftly implemented.

## Methods

### Clinical cohorts, study plan and ethical clearance

The present prospective study was conducted in the period between October 2020 and April 2021 (2^nd^ wave) in the Emergency Department at the Klinikum Hochrhein, Waldshut-Tiengen, Germany. Initially, an algorithm for the testing strategy and patient pathways was developed, established and implemented (Figure 1).

**Figure 1.**
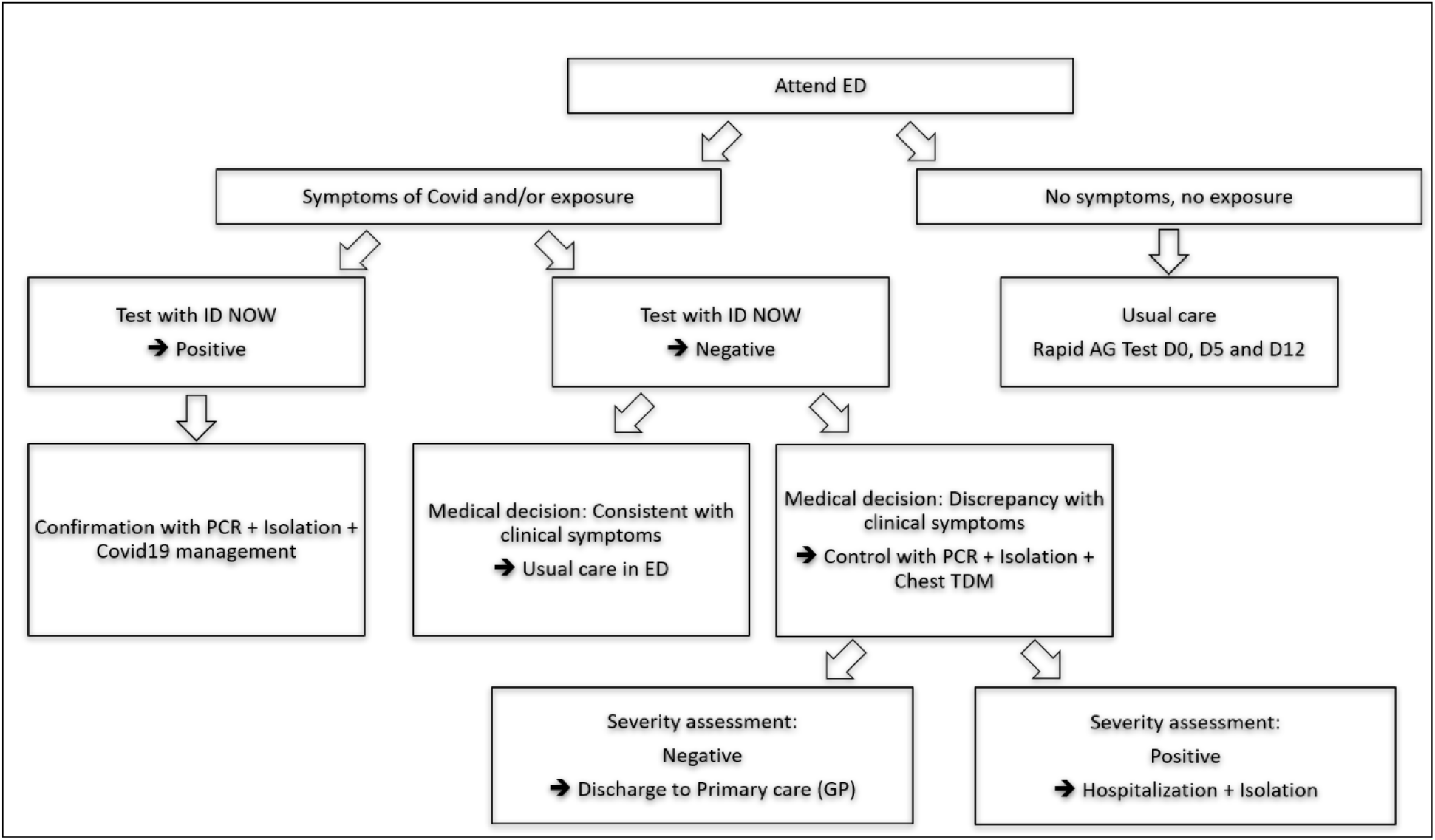
Testing Strategy and Patient Pathways

Patients aged 14-100 (median age 63) recruited into the study were included irrespective of the reason for referral to the emergency department but showed symptoms associated with COVID-19 and / or a high epidemiological risk (Table 1) [3, 13]. The evaluation of the collected findings within the scope of the study was carried out with the consent of the patients and completely anonymised.

**Table 1.**
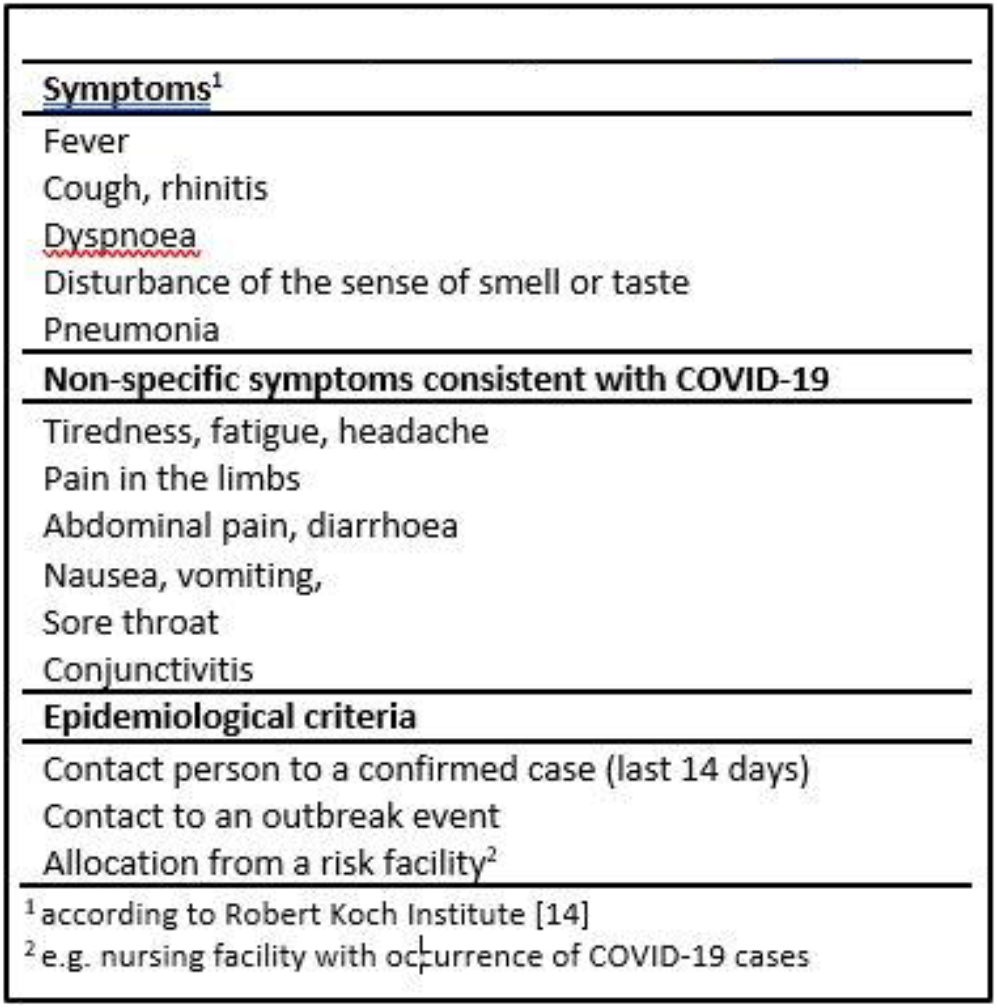
Criteria for performing the ID NOW™ <u>Test</u>

### Sample collection and testing methods

To obtain material for testing, the sterile foam swab supplied with the ID NOW™ COVID-19 test (Abbott, Chicago, USA) was used to collect anterior nasal specimens from patients arriving in the ambulance whilst in a prone position. The patients remained in the vehicle until the results were available, unless the individual required immediate vital care (e.g. such as shock rooms for heart attacks) and in those cases, treatment proceeded under full protective measures. For patients arriving on foot, a separate anteroom of the emergency department was used for initial diagnostics. Again anterior nasal swabs were collected with the provided sterile foam swab during an initial clinical assessment by a qualified nurse. All swabs were measured directly via the ID NOW™ COVID-19 test as point-of-care diagnostics in the emergency department without further processing or delay. Everyone that was allowed to perform the assays had been previously instructed on how to use the test by the manufacturer or by trained personnel. The tests were consistently carried out according to the instructions for use for the ID NOW™. Following a positive result with the ID NOW™ platform, a second nasopharyngeal swab (viscose tip, REF 09-812-8052, nerbe plus GmbH, Germany) was obtained within an hour from the patient for RT-PCR measurements (cobas SARS-COV-2 RT-PCR, Roche), dissolved in 3 ml virus transport medium and sent to the cooperating laboratory for analysis (Clotten Laborverwaltung GmbH, Freiburg). On average it took a minimum of 24 hours to obtain the results but in practice it was usually 36-48 hours.

### Statistical analysis

All test results as well as the clinical findings of the patients were documented in the electronic patient file of our hospital information system. After extraction and completely anonymised processing of the data, further statistical evaluations were carried out using SPSS v23 software. The performance data of the ID NOW™ COVID-19 test were calculated in comparison to RT-PCR as a reference. Deviating results were subjected to a clinical case-by-case examination and evaluated. In the case of positive results of the RT-PCR, the Ct values were additionally evaluated as a measure of the viral load present in the sample.

## Results

### ID NOW™ COVID-19 test shows high sensitivity in an ED setting

From a total of 710 symptomatic and true asymptomatic patients, 490 (69 %) were determined to be positive for SARS-CoV-2 using RT-PCR which included 472 individuals presenting symptoms or had been exposed and at least 18 asymptomatic (e.g. staff). The manufacturer’s information on the time required to obtain a result proved to be correct for negative results. For positive results, data were often available after just a few minutes (2 to 5) which further accelerated patient pathways. 488/490 (99.6%) of on-site tested samples were positive for the ID NOW™ COVID-19 test. The Ct values in the confirmatory RT-PCR provided an indication of the viral load which was between Ct values 7.94 and 37.42 (mean 24.41, Figure 2).

**Figure 2.**
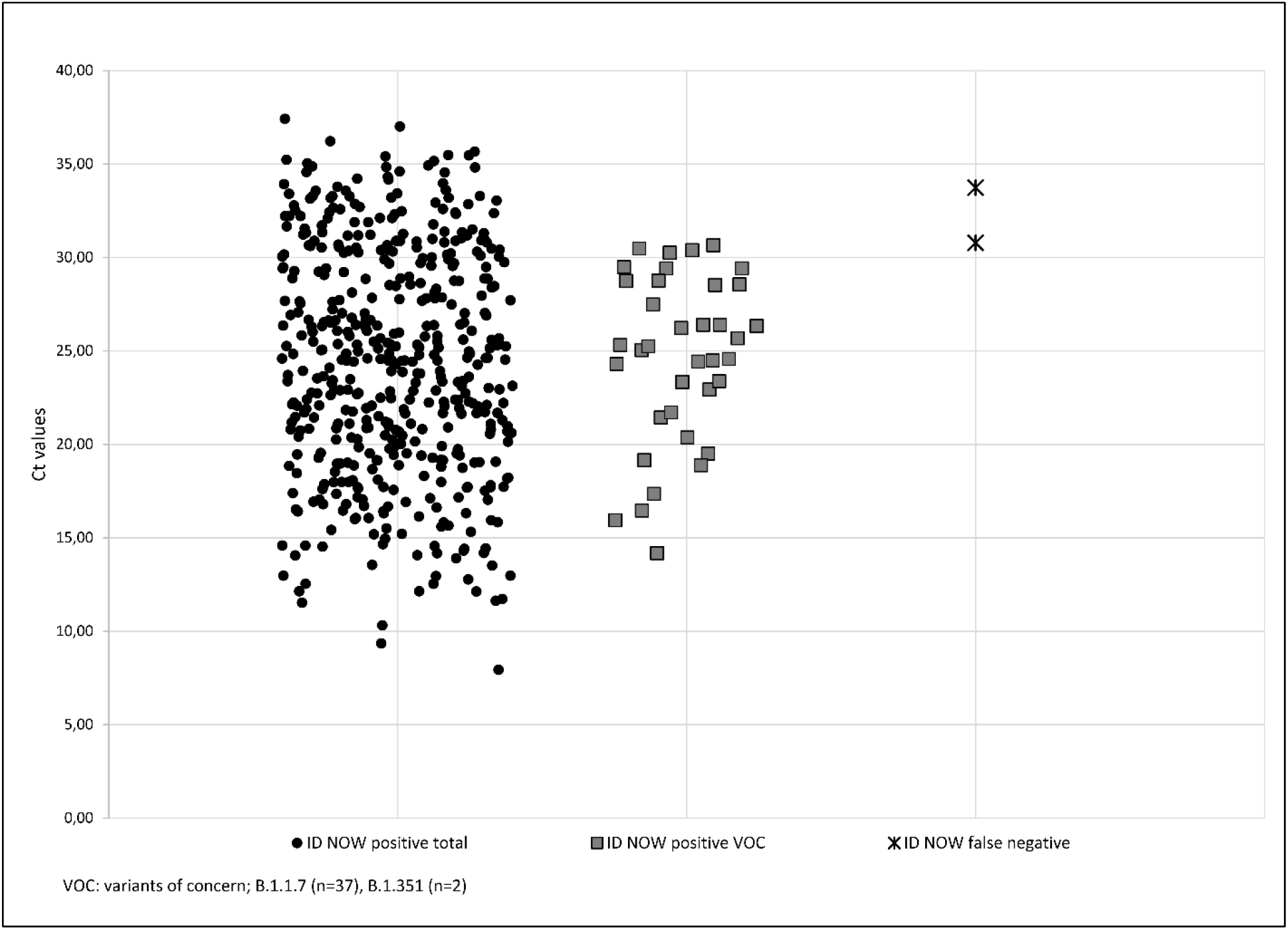
Comparison of Ct Values for Positive RT-PCR Results

In 113 samples (23.2%), the Ct value was above 30 and thus above the threshold specified by the Robert Koch Institute for possible de-isolation of the patients since the viral load in these samples was clearly below 1×10^6^ genome copies per ml [14].

During the study period, a total of 39 individuals presented infections with a variant of concern (VOC) and had corresponding Ct values between 14.18 and 30.66. Following genome sequencing it was determined that 37 cases presented B.1.1.7 (alpha variant) and 2 cases of B.1.351 (beta variant). All patients with confirmed VOC tested positive with the ID NOW™ COVID-19 test on arrival at the Emergency Department. 2 samples tested false negative with the ID NOW™ platform and on further evaluation, the corresponding Ct values determined in the RT-PCR were 30.78 and 33.73.

Thus, the overall diagnostic agreement of the test procedure was 98 % and applying Cohen’s Kappa as a measure of interrater reliability was calculated to be 0.95 (95 % CI 0.93 - 0.98) (Table 2).

**Table 2.** Performance data of ID NOW™ COVID-19 test with reference RT-PCR

| **Key Parameters** | **Value** | **95 % confidence interval** |
| --- | --- | --- |
| Sensitivity | 99.59 % | 98.53 – 99.95 |
| Specificity | 94.55 % | 90.67 – 97.15 |
| Positive predictive value | 97.60 % | 95.85 – 98.75 |
| Negative predictive value | 99.05 % | 96.60 – 99.88 |
| Diagnostic agreement | 0.980 |  |
| Positive percentage match | 0.996 | 0.985 – 1.000 |
| Negative percentage match | 0.945 | 0.907 – 0.972 |
| Cohen's Kappa | 0.95 | 0.93 – 0.98 |
| Positive diagnostic likelihood ratio | 18.26 | 10.53 – 31.65 |
| Negative diagnostic likelihood ratio | 0.004 | 0.001 – 0.017 |

According to Landis and Koch, this corresponds to an “almost perfect agreement” [14]. The sensitivity of the ID NOW™ COVID-19 test was 99.59 % (95 % CI 98.53 - 99.95), the specificity 94.55 % (90.67 - 97.15). The positive predictive value was calculated to be 97.6 % (95.85 - 98.75), the negative predictive value with 99.05 % (96.6 - 99.88). Likelihood ratios in evidence based medicine are a means to assess the value of a particular diagnostic test and use sensitivity and specificity to determine if a condition truly exists. In the settings here, the positive diagnostic likelihood ratio (DLR+) was 18.26 (10.53 - 31.65), and the negative likelihood ratio (DLR-) was 0.004 (0.001 - 0.017).

### Understanding false positive and false negative results when determining the clinical picture

14 cases from 710 patients presented discrepancies between results detected with the ID NOW™ COVID-19 assay performed directly in the Emergency Department and the RT-PCR performed in our collaborating laboratory (false negative 2 (0.28%); false positive 12 (1.69%)). These cases were then subjected to clinical and epidemiological case-by-case review and evaluation (Table 3).

**Table 3.** Clinical and epidemiological testing of discordant test results

|  |  |  |
| --- | --- | --- |
| **ID NOW™ false negative** |  |  |
| **No.** | **Facts** | **Evaluation** |
| 1 | Clinical-epidemiological<sup>1</sup> confirmed COVID-19; onset of disease > 10 days prior to admission and testing; AG rapid test negative. | COVID-19, late stage |
| 2 | Clinical-epidemiologically<sup>1</sup> confirmed COVID-19; detection via RT-PCR 25 days prior to admission and testing; AG rapid test negative. | COVID-19, late stage |
| **ID NOW™ false positive** |  |  |
| **No.** | **Facts** | **Evaluation** |
| 3 | Close contact with a person with a confirmed infection; tested negative on admission (incubation period); ID NOW™ positive 2 days after admission. | COVID-19, very early stage. |
| 4 | Close contact with a confirmed case (spouse); ID NOW™ positive on admission; AG rapid test negative. | COVID-19, very early stage. |
| 5 | Concomitant symptoms compatible with COVID-19; epidemiologic context unclear; AG rapid test negative. | Symptomatic, unexplained |
| 6 | Epidemiologically confirmed COVID-19; detection via RT-PCR approximately 28 days prior to admission and testing; AG rapid test negative. | State after Covid-19 |
| 7 | Clinically indubitable COVID-19; ID NOW™ and rapid AG test positive on admission; ill for several days; pneumogenic sepsis. | COVID-19, Advanced |
| 8 | Patients from a high-risk institution with an outbreak; AG rapid test negative; exact anamnesis not possible. | State after infection with few symptoms |
| 9 | Patients from a high-risk institution with an outbreak; AG rapid test negative; exact anamnesis not possible | State after infection with few symptoms |
| 10 | Asymptomatic; epidemiologic context unclear; AG rapid test negative. | Unsettled |
| 11 | Patients from a high-risk institution with an outbreak; AG rapid test negative; exact anamnesis not possible. | Infection with few symptoms |
| 12 | Anamnestic COVID-19 confirmed; detection via RT-PCR approximately 35 days prior to admission and testing; AG rapid test negative. | State after Covid-19 |
| 13 | Anamnestic COVID-19 confirmed; detection via RT-PCR approximately 30 days prior to admission and testing; AG rapid test negative. | State after Covid-19 |
| 14 | Anamnestic COVID-19 confirmed; detection via RT-PCR approximately 4 months prior to admission and testing; AG rapid test negative. | State after Covid-19 |
| AG: Antigen |  |  |
| <sup>1</sup> according to Robert Koch Institute [14] |  |  |

Both patients with a false negative ID NOW™ test were admitted to the hospital because of respiratory symptoms. In both cases, infection with SARS-CoV-2 had been confirmed by RT-PCR more than 10 days (patient 1) and 25 days (patient 2) before admission.

The RT-PCR performed on admission showed Ct values above 30 in each case as an indication of a low viral load in the smear material, which was related to the previously known, longer-standing infection. An assumed heterogeneity of the low viral load between the two smears could be an explanation as for the results. A supplementary antigen rapid test (SARS-COV-2 Rapid Antigen Test, REF 9901-NCOV-01G, Roche) carried out in parallel was negative in both cases.

Among the patients with a primary false positive result with the ID NOW™ COVID-19 test, 3 observations occurred repeatedly: i) a past infection with SARS-CoV-2 (n=4), ii) infections in the very early stage which were in close contact to confirmed cases (n=2) and iii) patients referred from a care facility that had had a significant outbreak but from which a reliable history was not possible (n=3). Patients referred from a nursing facility were hospitalised for reasons unrelated to COVID-19. It stands to reason that they had experienced low-symptoms and therefore infections had gone unnoticed. We hypothesise that these 9 patients also experienced random heterogeneities between smears at low viral loads.

One patient was admitted with a clinical, epidemiological and radiologically proven advanced COVID-19 disease state. The exact onset of symptoms could not be determined but the result of the ID NOW™ COVID-19 test as well as the rapid antigen test (SARS-COV-2 Rapid Antigen Test, REF 9901-NCOV-01G, Roche) were positive on admission. When the negative result of the RT-PCR arrived, the patient had already died due to multi-organ failure upon onset of pneumogene sepsis, so that a repeat examination could not be performed. It is possible that a low viral load was already present in this case, although errors in the pre-analysis cannot be ruled out. In 2 cases, reasons for discrepant examination results could not be conclusively clarified, of which 1 patient showed symptomatology compatible with COVID-19.

## Discussion

As an emergency department and primary point of contact for all emergency patients, rapid and reliable detection or exclusion of SARS-CoV-2 infection is imperative to ensure early and appropriate patient management and to prevent nosocomial spread of the infection. Confirmation of results by RT-PCR regularly took between 36-48 hours whereas the performance of the ID NOW™ COVID 19 test in the context of this study often required less than the stated 15 minutes. The test was carried out by trained nursing staff and integrated into the clinical process of admitting emergency patients, was well accepted by staff and considered “normal routine” after a few weeks demonstrating that the obtained results were reliable and trustworthy.

The performance data acquired during this study are excellent in terms of both positive and negative prediction compared to RT-PCR and show almost complete diagnostic agreement. The analysis of cases with discordant results suggests a connection with a very early or far advanced stage of infection with considerable fluctuations in the number of SARS-CoV-2 genome copies in the test material, whereby an influence on the results by pre-analytical factors cannot be completely ruled out either. Both cases, initially classified as “false negative”, showed Ct values >30 in RT-PCR, which corresponds to a viral load clearly below 1×10^6^ genome copies per ml [13]. In accordance with current publications, we assume that these patients can be considered as not (anymore) infectious at the time of testing [15-18]. In addition, 18 asymptomatic cases were caught by the ID NOW™ platform; these were random individuals and staff members that had had a positive antigen test or been in close contact with a positive case. All cases were confirmed positive through an additional PCR test. Moreover, individuals that initially tested negative in this study but remained in the hospital were also negative 5 or 12 days later since control testing was carried out with further ID NOW™ COVID-19 tests. Even though no significance can be deduced from these observations it clearly indicates the reliability of the ID NOW™ system even in asymptomatic persons.

In contrast to the majority of previously published studies, the data presented here were obtained under real clinical conditions within the framework of a structured algorithm and in strict compliance with the manufacturer’s instructions. Here there was no dissolution of the specimen in transport medium and/or delayed analysis (≥ 2 h), which have been practised elsewhere and ultimately represent an “off-label use”. Such methods have been shown to significantly impair the reliability of the method [7, 11, 19-23]. A recent study from Nguyen Van and colleagues proved this in a real-life setting in an ED: placing specimens in transport medium and testing hours later reduced the sensitivity of the test by 35.5% [2]. As with all molecular biological tests for SARS-CoV-2, the results must always be evaluated in the context of the clinical picture and epidemiological history of the patient in order to arrive at a robust clinical decision. Taking this into account, results obtained from the ID NOW™ COVID-19 test and RT-PCR should be considered equivalent. Whereby RT-PCR requires considerably more time until the results are available, the ID NOW™ COVID-19 test provides initial data within 15 minutes. The ID NOW™ COVID-19 test also fulfils the requirements of the official case definition by the Robert Koch Institute [24], so that RT-PCR is dispensable as a pure confirmatory test.

The testing strategy that was applied during this study in the second wave required specimens to be collected in the ambulance or a separate room for walk-ins. Following a brief training of staff on using the device testing ran smoothly in day to day operations. Moreover, the implementation of the test in our clinical processes consistently allowed the correct classification of patients within minutes, allowed immediate decisions regarding whether to isolate or cohort individuals alongside separating patients’ pathways. Furthermore, later in the course of the disease, a decision to remove isolation and discharge to a care facility could be based on ID NOW™ COVID -19 testing. This was in drastic contrast to the process during the first wave were RT-PCR results could take up to 48 hours. During this time, the positive patient had to be isolated in a separate room which impacted the hospital further since patients were then turned away due to lack of space and moreover, patients in urgent need of isolation could not be admitted. It is estimated therefore that adding the ID NOW™ platform not only accelerated patient placement, care and therapy but reduced hospital costs by 300-400 Euro (price of room blocking for 24 hours) per suspected patient. Thus, it is anticipated that this reliable point-of-care testing will result in an overall economic benefit through the avoidance of bed blocks due to individual isolations and more targeted use of inpatient resources, which would have to be quantified in further studies.

The present data stem from a single-centre evaluation of the ID NOW™ COVID-19 test for SARS-CoV-2 in emergency patients. Transferability to other patient groups and departments needs to be assessed. With regard to protection against nosocomial spread of infection, the ID NOW™ COVID-19 test, like any other diagnostic test procedure, may have a “blind spot” when the patient is admitted to the hospital for other reasons and is still in the incubation phase at that time. We therefore consider it useful to repeat the test on the 3^rd^ and 5^th^ day of the inpatient stay in non-isolated patients and as commented above, none of the originally negative patients became positive on days 3 and 5. Moreover, the analysis of discordant results in our study shows that each test resulting in a positive SARS-CoV-2 outcome must always be evaluated in the context of clinical symptoms and anamnesis, regardless of the method. In particular, it must be taken into account that in very early and very late phases of infection, the viral load in the smear material is subject to strong fluctuations.

The ID NOW™ COVID-19 test is a safe molecular method in the primary diagnosis of an infection with SARS-CoV-2 and for the management of clinical processes, even taking into account the official case definition. However, a supplementary RT-PCR is required if a Ct value or the detection of a viral variant appears to be significant in a specific case. Ultimately, the concrete benefit of point-of-care diagnostics depends largely on the organisation of the emergency department and the deployment of trained staff but during the second wave of infection proved invaluable in stream-lining patient pathways, immediate therapy and cost-effectiveness.

## Conclusions

In view of the high number of patients and the resulting pressure on the healthcare system, there is an urgent need for user-friendly, fast and safe point-of-care diagnostics. The present prospective evaluation of the ID NOW™ COVID -19 test for SARS-CoV-2 in a real clinical context within an emergency department shows that this platform provide a very fast and reliable alternative to a laboratory-based RT-PCR and enables appropriate patient management already at the time of admission and the avoidance of nosocomial infections. Through targeted use of the hospital’s inpatient resources, the impact of the pandemic could be mitigated and standard care largely maintained.

## Data Availability

All data are archived and available with the authors

## Acknowledgements

The authors send their sincere thanks to the participants of the study project and to all staff members who were involved in specimen handling and testing; their efforts were greatly appreciated.

## Literature

1. Collier DA, Assennato SM, Warne B et al (2020) Point of Care Nucleic Acid Testing for SARS-CoV-2 in Hospitalized Patients: A Clinical Validation Trial and Implementation Study. Cell Rep Med 1(5):100062. doi:10.1016/j.xcrm.2020.100062

2. Nguyen Van J-C, Gerlier C, Pilmis B, Mizrahi A, Péan de Ponfilly G, Khaterchi A, Enouf V, Ganansia O, Le Monnier A (2021) Prospective evaluation of ID NOW COVID-19 assay used as point-of-care test in an Emergency Department.

3. Kortüm S, Becker D, Ott H-J, Schlaudt H-P (2020) The Role of the Emergency Department in Protecting the Hospital as a Critical Infrastructure in the Corona Pandemic Strategies and Experiences of a Rural Sole Acute-Care Clinic. https://doi.org/10.1101/2020.09.07.20185819

4. Abbott Diagnostics Scarborough, Inc. (2020) ID NOW™ COVID-19 Package Insert. https://www.globalpointofcare.abbott/en/product-details/id-now-covid-19.html. Accessed: 13 May 2021

5. Harrington A, Cox B, Snowdon J, Bakst J, Ley E, Grajales P, Maggiore J, Kahn S (2020) Comparison of Abbott ID Now and Abbott m2000 Methods for the Detection of SARS-CoV-2 from Nasopharyngeal and Nasal Swabs from Symptomatic Patients. J Clin Microbiol 58(8). doi:10.1128/JCM.00798-20

6. Rhoads DD, Cherian SS, Roman K, Stempak LM, Schmotzer CL, Sadri N (2020) Comparison of Abbott ID Now, DiaSorin Simplexa, and CDC FDA Emergency Use Authorization Methods for the Detection of SARS-CoV-2 from Nasopharyngeal and Nasal Swabs from Individuals Diagnosed with COVID-19. J Clin Microbiol 58(8). doi:10.1128/JCM.00760-20.

7. Zhen W, Smith E, Manji R, Schron D, Berry GJ (2020) Clinical Evaluation of Three Sample-to-Answer Platforms for Detection of SARS-CoV-2. J Clin Microbiol 58(8). doi:10.1128/JCM.00783-20

8. Dhamad AE, Abdal Rhida MA (2020) COVID-19: molecular and serological detection methods. PeerJ 8:e10180. doi:10.7717/peerj.10180

9. Subsoontorn P, Lohitnavy M, Kongkaew C (2020) The diagnostic accuracy of isothermal nucleic acid point-of-care tests for human coronaviruses: A systematic review and meta-analysis. Sci Rep 10(1):22349. doi:10.1038/s41598-020-79237-7.

10. Farfour E, Asso-Bonnet M, Vasse M (2021) The ID NOW COVID-19, a high-speed high-performance assay. Eur J Clin Microbiol Infect Dis. 15 : 1–5. doi: 10.1007/s10096-021-04243-0.

11. Khan P, Aufdembrink LM, Engelhart AE (2020) Isothermal SARS-CoV-2 Diagnostics: Tools for Enabling Distributed Pandemic Testing as a Means of Supporting Safe Reopenings. ACS Synth Biol. doi:10.1021/acssynbio.0c00359

12. Basu A, Zinger T, Inglima K, Woo K-M, Atie O, Yurasits L, See B, Aguero-Rosenfeld ME (2020) Performance of Abbott ID Now COVID-19 Rapid Nucleic Acid Amplification Test Using Nasopharyngeal Swabs Transported in Viral Transport Media and Dry Nasal Swabs in a New York City Academic Institution. J Clin Microbiol 58(8). doi:10.1128/JCM.01136-20

13. Stokes W, Berenger BM, Singh T, Adeghe I, Schneider A, Portnoy D, King T, Scott B, Pabbaraju K, Shokoples S, Wong AA, Gill K, Turnbull L, Hu J, Tipples G (2020) Acceptable Performance of the Abbott ID NOW Among Symptomatic Individuals with Confirmed COVID-19.

14. Robert Koch Institute (2021) Epidemiological profile of SARS-CoV-2 and COVID-19. Status 19.4.2021. https://www.rki.de/DE/Content/InfAZ/N/Neuartiges_Coronavirus/Steckbrief.html. Accessed: 16 May 2021

15. Landis JR, Koch GG (1977) The measurement of observer agreement for categorical data. Biometrics 33(1):159–174

16. van Kampen JJA, van Vijver DAMC de, Fraaij PLA, Haagmans BL, Lamers MM, Okba N, van Akker JPC den, Endeman H, Gommers DAMPJ, Cornelissen JJ, Hoek RAS, van der Eerden MM, Hesselink DA, Metselaar HJ, Verbon A, Steenwinkel JEM de, Aron GI, van Gorp ECM, van Boheemen S, Voermans JC, Boucher CAB, Molenkamp R, Koopmans MPG, Geurtsvankessel C, van der Eijk AA (2021) Duration and key determinants of infectious virus shedding in hospitalized patients with coronavirus disease-2019 (COVID-19). Nature Communications 12(1):1–6. doi:10.1038/s41467-020-20568-4

17. Bullard J, Dust K, Funk D, Strong JE, Alexander D, Garnett L, Boodman C, Bello A, Hedley A, Schiffman Z, Doan K, Bastien N, Li Y, van Caeseele PG, Poliquin G (2020) Predicting infectious SARS-CoV-2 from diagnostic samples. Clin Infect Dis. doi:10.1093/cid/ciaa638

18. Wölfel R, Corman VM, Guggemos W, Seilmaier M, Zange S, Müller MA, Niemeyer D, Jones TC, Vollmar P, Rothe C, Hoelscher M, Bleicker T, Brünink S, Schneider J, Ehmann R, Zwirglmaier K, Drosten C, Wendtner C (2020) Virological assessment of hospitalized patients with COVID-2019. Nature <London> 581(7809):465–469. doi:10.1038/s41586-020-2196-x

19. Krause E, Puyskens A, Bourquain D, Brinkmann A, Biere B, Schaade L, Michel J, Nitsche A (2021) Sensitive on-site detection of SARS-CoV-2 by ID NOW COVID-19.

20. Procop GW, Brock JE, Reineks EZ, Shrestha NK, Demkowicz R, Cook E, Ababneh E, Harrington SM (2021) A Comparison of Five SARS-CoV-2 Molecular Assays With Clinical Correlations. Am J Clin Pathol 155(1):69–78. doi:10.1093/ajcp/aqaa181

21. Panpradist N, Wang Q, Ruth PS, Kotnik JH, Oreskovic AK, Miller A, Stewart SW, Vrana J, Han PD, Beck IA, Starita LM, Frenkel LM, Lutz BR (2021) Simpler and faster Covid-19 testing: strategies to streamline SARS-CoV-2 molecular assays. EBioMedicine 64. doi:10.1016/j.ebiom.2021.103236

22. Mahmoud SA, Ibrahim E, Ganesan S, Thakre B, Teddy JG, Raheja P, Zaher WA (2021) Evaluation of seven different rapid methods for nucleic acid detection of SARS-COV-2 virus.

23. Lephart PR, Bachman MA, LeBar W, McClellan S, Barron K, Schroeder L, Newton DW (2021) Comparative study of four SARS-CoV-2 Nucleic Acid Amplification Test (NAAT) platforms demonstrates that ID NOW performance is impaired substantially by patient and specimen type. Diagn Microbiol Infect Dis 99(1):115200. doi:10.1016/j.diagmicrobio.2020.115200

24. Farfour E, Roux A, Ballester M, Gagneur L, Renaux C, Jolly E, Vasse M (2020) Improved performances of the second generation of the ID NOW influenza A&B 2® and comparison with the GeneXpert®. Eur J Clin Microbiol Infect Dis 39(9):1681–1686. doi:10.1007/s10096-020-03905-9

25. Robert Koch Institute (2020) Case definition Coronavirus Disease 2019 (COVID-19) (SARS-CoV-2), as of 23.12.2020. https://www.rki.de/DE/Content/InfAZ/N/Neuartiges_Coronavirus/Falldefinition.pdf?blob=publicationFile. Accessed: 09 May 2021

